# Data-driven prediction of spinal cord injury recovery: an exploration of current status and future perspectives

**DOI:** 10.1101/2024.05.03.24306807

**Authors:** Samuel Håkansson, Miklovana Tuci, Marc Bolliger, Armin Curt, Catherine R. Jutzeler, Sarah Brüningk

## Abstract

Spinal Cord Injury (SCI) presents a significant challenge in rehabilitation medicine, with recovery outcomes varying widely among individuals. Machine learning (ML) is a promising approach to enhance the prediction of recovery trajectories, but its integration into clinical practice requires a thorough understanding of its efficacy and applicability. We systematically reviewed the current literature on data-driven models of SCI recovery prediction. The included studies were evaluated based on a range of criteria assessing the approach, implementation, input data preferences, and the clinical outcomes aimed to forecast. We observe a tendency to utilize routinely acquired data, such as International Standards for Neurological Classification of SCI (ISNCSCI), imaging, and demographics, for the prediction of functional outcomes derived from the Spinal Cord Independence Measure (SCIM) III and Functional Independence Measure (FIM) scores with a focus on motor ability. Although there has been an increasing interest in data-driven studies over time, traditional machine learning architectures, such as linear regression and tree-based approaches, remained the overwhelmingly popular choices for implementation. This implies ample opportunities for exploring architectures addressing the challenges of predicting SCI recovery, including techniques for learning from limited longitudinal data, improving generalizability, and enhancing reproducibility. We conclude with a perspective, highlighting possible future directions for data-driven SCI recovery prediction and drawing parallels to other application fields in terms of diverse data types (imaging, tabular, sequential, multimodal), data challenges (limited, missing, longitudinal data), and algorithmic needs (causal inference, robustness).

## Introduction

Spinal Cord Injury (SCI) is a complex condition that presents a significant challenge in medical rehabilitation, as it involves a range of physiological response cascades. Beyond the initial physical trauma (e.g. comprising fractures and axonal disruption), the physiological and mechanical damage to spinal tissues triggers neuronal excitotoxicity and increases reactive oxygen concentrations and glutamate levels [1], resulting in neurological dysfunction [2]. As a result, the clinical presentation exhibits extensive heterogeneity in terms of motor abilities (e.g. walking, grasping), sensory function, and various aspects of functionality, including sexual, bowel, and bladder functions. Given the substantial heterogeneity in terms of injury characteristics, clinical presentation, and recovery of bodily functions [3], predicting outcomes at the individual level is challenging. Achieving accurate recovery prediction is, however, essential for establishing realistic expectations among patients and their families regarding long-term prognosis [4]. Moreover, accurate and reliable outcome prediction is crucial for designing and implementing effective clinical trials.

Given the increasing availability of large SCI databases [5–11], it is a logical step to consider data-driven models for recovery prediction. Based on parameterization, these models allow for the establishment of a connection between (multi-modal) input data and outcomes to facilitate predictions. However, it is essential to carefully choose the right model architecture in light of available training data, sample sizes, and clinical questions of interest. For successful clinical deployment, a model must satisfy stringent expectations regarding robustness, generalizability, interpretability, and reproducibility, in addition to meeting performance criteria. Robustness refers to the ability to preserve performance despite conditions such as noisy data and distribution shifts. In general, this requires learning relevant relationships between input data and output labels, rather than overfitting the model to the training data. Generalizability conveys that the method has a high sample size-to-complexity ratio and is externally validated meaning that it performs well on new data, from the same and other origins. In the clinical application context, tracing the prediction’s underlying process is essential to building trust in the model and validating that the decision process is well-founded. Interpretability and explainability analysis facilitate this [12]. Finally, the research and implementation should also be reproducible, meaning that the methods, data processing, and input and output data are transparent and readily usable by others.

This review aims to provide a comprehensive overview of the current state of data-driven applications in the context of SCI recovery prediction. We begin by systematically exploring the diverse landscape within this field, summarizing the various types of input data, outcomes, prediction models, and evaluation methods reported. Transitioning from discussing the status quo and state-of-the-art in the SCI field today, we provide a perspective regarding promising future directions and inherent challenges. In particular, we stress examples and parallels drawn from other data science applications in healthcare and discuss how concepts and architectural designs from these domains could be adapted for SCI-related tasks.

## Methods

### Search strategy

We conducted a systematic search across three major bibliographic databases: PubMed, Scopus, and Web of Science. Included in our search were studies published from the inception of these databases up to October 2nd, 2023 (search date). Our search strategy comprised a carefully constructed query for: (“machine learning” OR “statistical model” OR “artificial intelligence” OR “deep learning” OR “decision tree” OR “random forest” OR “prediction rule” OR “prediction model”) AND (“spinal cord injury” OR “spinal cord lesion”) AND (recovery OR prognosis OR prediction OR outcome). Our search was not restricted by language. Additionally, manual searching was also performed, reviewing reference lists of relevant articles and comprehensive reviews.

### Inclusion and exclusion criteria

We included peer-reviewed articles focusing on predicting recovery in human adults following acute SCI. Our definition of “recovery” entails predicting motor, sensory, functional, or other clinically important outcomes at a later stage from the acute injury phase. No specific restrictions were imposed regarding the longitudinal timeframe for inclusion. We excluded articles involving pediatric patients or non-human subjects, and those related to non-traumatic or non-ischemic spinal cord injury. Additionally, studies predicting associated or secondary conditions post-injury (e.g. pressure ulcers, surgery-related complications) and those focusing on non-recovery outcomes (like survival, hospital stay length) or investigative treatment effects were excluded. Duplicate studies, abstracts, systematic reviews, meta-analyses, case studies, and validation studies of models already included in this review were also not considered. Reasons for exclusion of studies were defined according to the hierarchy in Table S1.

### Selection process

We assessed the suitability for inclusion by reviewing titles and abstracts, followed by full-text. This assessment was independently conducted by two reviewers (MT and SH). Conflicting opinions regarding study inclusion or exclusion were settled on a case-by-case basis by mutual discussion with additional investigators (CJ, SB).

### Data extraction

Two independent reviewers (MT, SH) performed the data extraction, which was initiated with the systematic retrieval of bibliographic information such as authorship details, publication years, publication titles, and Digital Object Identifiers (DOI).

From full texts, we gathered detailed data including: (i) the types of prediction models, (ii) outcomes, (iii) input data, (iv) information about the model architecture(s), implementation, and training, and (v) model performance and metrics used for evaluation.

#### Qualitative assessment

We evaluated the quality of the included studies based on different criteria spanning five categories: *Clinical Significance*, *Reproducibility*, *Generalizability*, *Machine Learning Quality*, and *Machine Learning Performance*. Each category received a rating from 0 (poor quality) to 5 (high quality) to indicate the quality of the study for the relevant criterion.

To evaluate the *Clinical Significance* of the included studies, we assigned a score of 5 to papers that clearly outlined the clinical problem addressed, employed pertinent outcome measures, and emphasized clinical impact. Conversely, studies lacking a clear motivation for the clinical application of the presented approach, such as purely computational studies, were assigned a score of 0.

*Reproducibility* refers to the ability to replicate the findings of a study by following the outlined methods as well as using the provided data and code. Ideally, a study provides clear input and output variables, a description of variable coding for input (binary, categorical, or numerical) and output (binary or multi-class classification, or regression), and an overview of the employed model architecture in addition to details regarding its implementation. Both model code (e.g. as a public repository) and supporting data should be available for external applications. We subtracted scoring points for any missing information.

Understanding the extent to which findings and data-driven models can be generalized across different patient populations, injury severities, or clinical settings is key to examining the potential for clinical deployment. Thus, we assessed the *Generalizability* of the selected studies as follows: a score of 5 was assigned if the risk of overfitting to training data was explicitly addressed. This was considered accomplished through either external validation or by performing a clean data split (with no apparent information leakage) between the training and test set. To achieve an optimal score, we further considered the model complexity with respect to the sample size, and if the utilized data were multi-institutional. We scored 0 when the model was deemed susceptible to overfitting.

Under the umbrella of *Machine Learning Quality*, aspects regarding model architecture, training, and evaluation choices were assessed. We assigned a *Machine Learning Quality* score of 5 to papers that comprehensively reported their entire training pipeline, addressed issues of overfitting and imbalanced data, included strategies for feature selection (where appropriate), and chose relevant evaluation metrics. Ideally, a benchmark of different architectures was performed. We scored a 0 if none of the above criteria were met.

*Machine Learning Performance* quantifies how well a model accomplishes the task it was designed for. Given the strong effect of the *Machine Learning Quality* on the *Performance*, we fixed the *Machine Learning Quality* score as the upper limit for the performance score. Excellent performance was quantified as an accuracy (or ROCAUC) exceeding 90% or a correlation coefficient of 0.8 while accounting for possible class imbalance. We assigned a score of 0 to articles when the prediction quality was not assessed or did not exceed random prediction. Intermediate scores were given depending on the ratio between excellent and achieved performance.

### Statistical analysis

Based on the above scoring metrics, we summarize the included studies by the evaluation criteria using boxplots. Moreover, we identified groups of studies by hierarchical clustering. Clustering was performed such that the cophenetic distance (the height of the dendrogram at the point where the two points are first joined together), between two observations is minimal, with a maximum of 6 flat clusters. The Python library SciPy [13] with the function fcluster in the subpackage cluster.hierarchy was used to generate the clusters and the hierarchical linkage was generated using linkage from the same subpackage.

## Results

### Study selection

Figure 1 shows a Consort diagram leading to the final set of included studies. Our search identified 973 studies, after the removal of 318 duplicates, 655 article abstracts were screened, resulting in the exclusion of 572 articles. The subsequent review of 83 full-text articles resulted in the exclusion of 19 more articles. Finally, 64 studies met our inclusion criteria. Moreover, one article was added outside of the literature search through the references in one of the included papers (a validation study was exchanged for the study where the method was first suggested), and one article was added manually, resulting in 66 manuscripts [14–79].

**Fig. 1.**
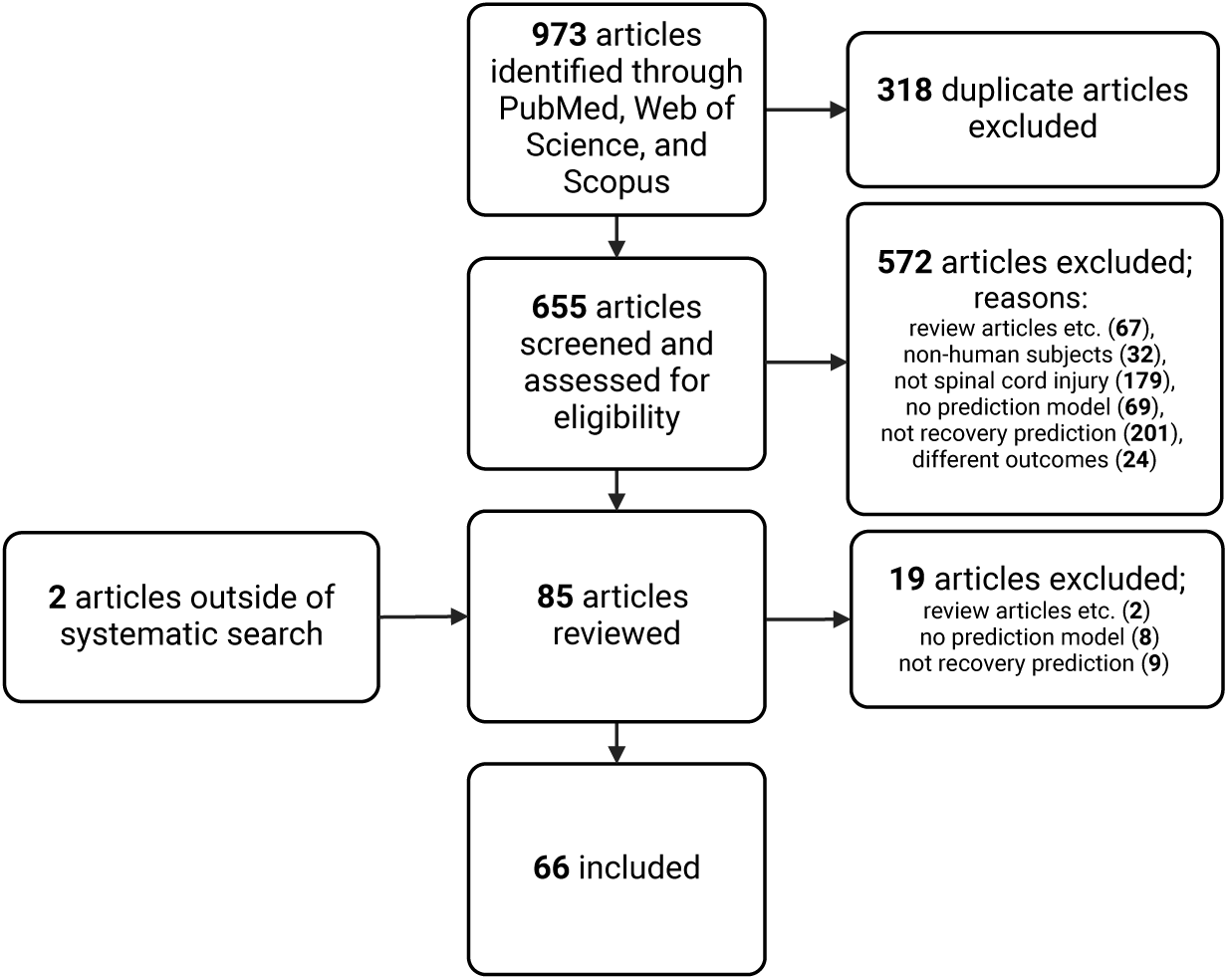
Flowchart representing the search strategy, retained studies, and exclusion contributions.

### Study characteristics

An overview of the extracted information from all included studies is available in supplementary Table S2.

#### Input and output data

A range of input and output data types were addressed in the covered studies as shown in Table 1 and Table 2. Patient demographics and injury severity, as measured by the American Spinal Injury Association (ASIA) Impairment Scale (AIS), were identified as primary input factors in 56 (85%) and 47 (71%) papers, respectively.

**Table 1.** Overview of Input data types. Note that multiple labels can be assigned to one study. NLI: Neurological Level of Injury. AIS: American Spinal Injury Association Impairment Scale. ISNCSCI: International Standards for Neurological Classification of SCI. UEMS: Upper Extremity Motor Score. LEMS: Lower Extremity Motor Score. TMS: Total Motor Score. VAC: Voluntary Anal Contraction. DAP: Deep Anal Pressure. FIM: Functional independence motor score. MRC: Manual Muscle Test. GRASSP: Graded and Redefined Assessment of Strength, Sensibility and Prehension. MP: Motor Point mapping.

| Category | Input Data types | N of Papers | % |
| --- | --- | --- | --- |
| Neurological | AIS | 47 | 71.2% |
|  | NLI | 34 | 51.5% |
|  | ISNCSCI | 21 | 31.8% |
|  | UEMS/LEMS/TMS | 17 | 25.8% |
|  | VAC/DAP | 1 | 1.5% |
|  | Pin prick | 9 | 13.6% |
| Functional | FIM | 12 | 18.2% |
|  | MRC | 5 | 7.6% |
|  | GRASSP/MP | 2 | 3.0% |
| Imaging | MRI features | 12 | 18.2% |
|  | MRI images | 4 | 6.1% |
| Other | Demographics | 56 | 84.8% |
|  | Comorbidities | 14 | 21.2% |
|  | CSF/Blood | 8 | 12.1% |

**Table 2.** Overview of output data types. Note that multiple labels can be assigned to one study. SCIM III: Spinal Cord Independence Measure. *Contracture: Spasticty and Contracture. **Bone hearling: Odontoid fracture nonunion, Bone Mineral Density and Bone Mineral Content. *** Unspecified: E.g. Soft tissues, Short Form 12-Questionnaire Health Survey.

| Category | Output Data types | N of Papers | % |
| --- | --- | --- | --- |
| Neurological | AIS | 21 | 31.8% |
|  | ISNCSCI | 5 | 7.6% |
|  | UEMS/LEMS/TMS | 5 | 7.6% |
|  | Contracture* | 2 | 3.0% |
|  | Pain type | 1 | 1.5% |
| Functional | Walking | 11 | 16.7% |
|  | SCIM III | 10 | 15.2% |
|  | FIM | 9 | 13.6% |
|  | Bowel/Bladder | 5 | 7.6% |
|  | Prehension | 1 | 1.5% |
| Other | Discharge destination | 1 | 1.5% |
|  | Bone healing** | 2 | 3.0% |
|  | Unspecified*** | 2 | 3.0% |

These data types were followed by the neurological level of injury (NLI, 51.5%); subsets of the ISNCSCI (e.g. pin prick scores, or selected myomotes, 31.8%); and the sum scores for upper extremity (UEMS), lower extremity (LEMS), or total motor scores (TMS) in 17 papers (25.8%).

Imaging data was also a popular input, appearing in 16 papers. “Imaging data” here refers to relevant modalities (e.g. MRI), but also extracted information from images, such as Brain and Spinal Injury Center (BASIC) scores [80]. Only two papers used algorithmic methods to process images rather than pre-extracted image features. Functional input data types included the Functional Independence Measure (FIM, 18.2%), Manual Muscle Tests (MRC, 7.6%), and the Graded Redefined Assessment of Strength, Sensibility, and Prehension (GRASSP, 3.0%).

Eight studies used molecular or hematological information such as blood markers [22] or proteomic analysis of cerebrospinal fluid (CSF) [71].

The specific endpoints addressed covered a variety of scenarios as shown in Table 2. Primarily, functional and neurological outcomes were predicted. The summarized functional endpoints consist of Spinal Cord Independence Measure (SCIM) III [81] (15.2%), 6-minute walk test [82] (16.7%), Functional Independence Measure (FIM) [83] (13.6%), Prehension Performance [84] (1.5%), bladder function (e.g. [81]) (4.6%), and bowel function (e.g. [81]) (3.1%). 34 out of 66 studies predicted neurological outputs, such as AIS grade conversion (31.8%), composite motor scores (LEMS, UEMS or TMS, 7.6%), 2-motor level improvement on the cervical myotomes on either left or right side [46] (1.5%), Percent Deficit Improvement [28] (PDI, 1.5%), which quantifies neurological recovery as the percentage improvement in motor, pin, and touch scores relative to the maximum possible improvement from the baseline to the endpoint score, and pin prick (1.5%) [66].

Moreover, the timing of the data acquisition and prediction endpoints relative to the time of injury were recorded (Table 3). Multiple studies (34) with clearly defined time points since injury considered input data at a maximum of 56 days (i.e. 8 weeks) after injury. One study used input data from between 3 weeks and 3 months [68]. In the remaining studies, the specific timing of input data acquisition was either unclear (22 studies) or not reported (9 studies). For outcomes, 21 out of 42 studies with reported time points predicted at up to 1 year post-injury. Among these, 12 studies reported 6 months as their prediction time, and 3 studies used an evaluation at 3 months post-injury.

**Table 3.** Summary of the time points for included input data and evaluation endpoints. Unspecific input time: admission time, acute phase, baseline, and rehab admission. Unspecific output time: at discharge (unknown if this refers to the primary care or rehabilitation facility), after 36 training sessions (unknown absolute time since injury), rehabilitation discharge (unknown absolute time since injury).

| Prediction Time | N papers |
| --- | --- |
| > 1 year | 6 |
| ~1 year | 21 |
| ~6 months | 12 |
| ~3 months | 3 |
| Unspecific | 17 |
| Not reported | 7 |
| Inputs | N papers |
| < 72 h | 6 |
| < 31 days | 19 |
| < 56 days | 9 |
| 3 weeks - 3 months | 1 |
| Unspecific | 22 |
| Not reported | 9 |

Moreover, six studies reported their prediction time to be *>* 1 year and two of these studies predicted outcomes at multiple times, *∼*1st year, *∼*2nd year, or *∼*5th year [52, 64]. Similar to input data, several studies (n=17) did not specify exact time stamps, but defined categories such as: “at discharge”, or the number of training sessions [54]. Seven studies did not report their prediction time, impeding reproducibility.

#### Prediction model architecture choices

The included studies encompass various computational architectures, ranging from traditional statistical models to advanced designs incorporating model ensembles and deep learning frameworks. Linear and logistic regression are recurrent choices (50.7%), leveraging their interpretability and user-friendly implementation. Tree-based models and ensembles, including random forest or extreme gradient boosting (XGBoost), were also popular (29%). This showcases the flexibility of tree-based structures to capture complex relationships within the data while being easily implementable given established libraries (e.g. using *scikit-learn* library [85]). Deep learning techniques such as neural networks were reported (9%) but the deliberate motivation for the choice of more complex architectures was rarely reported. Figure 2C summarizes the distribution of the different model architectures over the time of publication. We observed a trend toward alternatives to linear models in recent years.

**Fig. 2.**
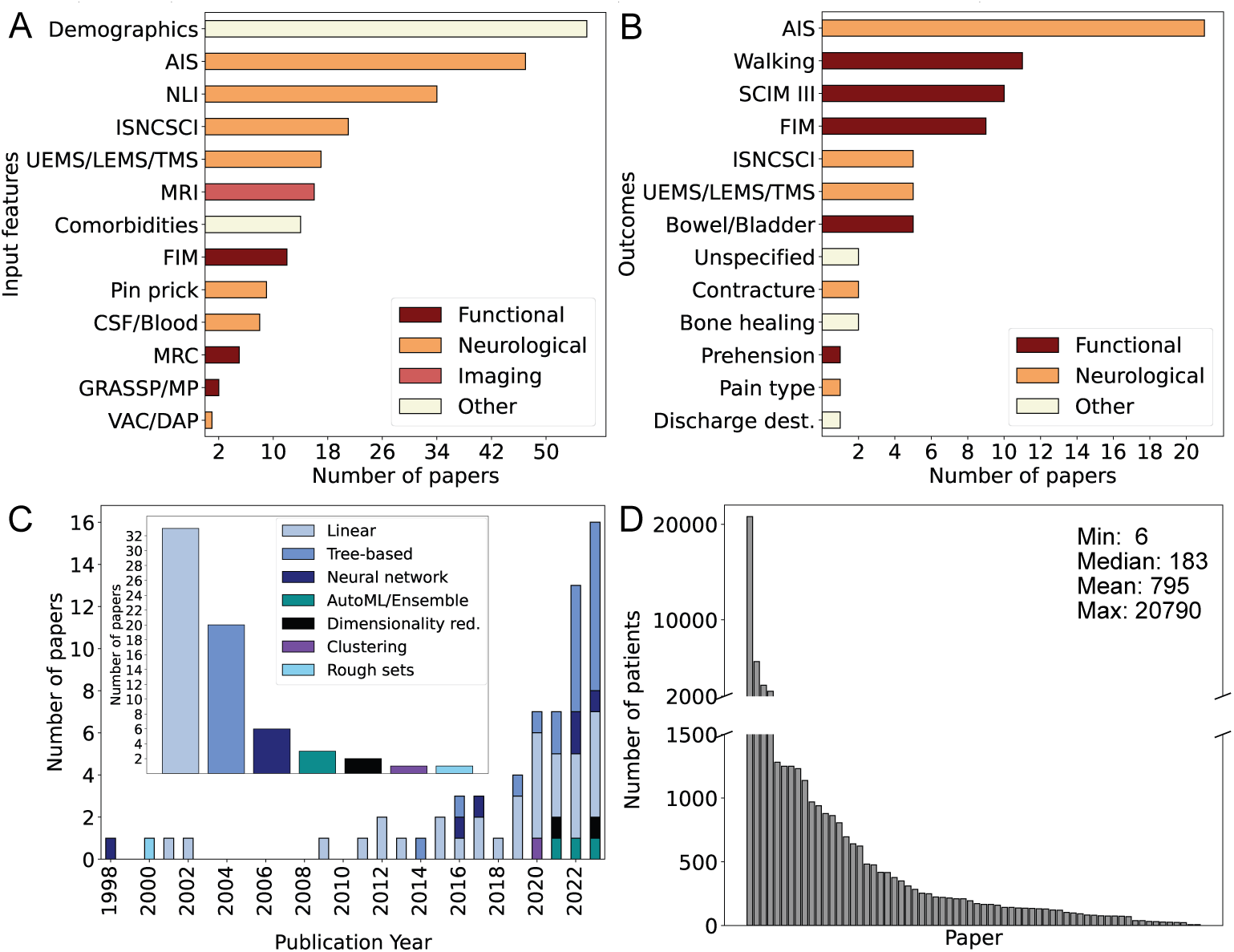
Overview of the models, inputs, outcomes, and number of patients used in the selected papers. **A) Input features**. Demographic data was used in almost all papers. AIS grade, neuro-logical level of injury (NLI), and neurological assessment data were common. Note that each study could utilize multiple outcomes and input features. **B) Outcomes**. AIS grade was the most common outcome followed by walking ability, FIM, and SCIM III. “Bone healing” refers to bone density and nonunion odontoid fractures. Additionally, “Bowel/Bladder” covers bowel management, bladder recovery, urinary continence, and urodynamic risk factors for upper urinary tract damage. **C) Number of publications per year**. The outer plot shows the increase in publications regarding recovery prediction models in SCI as well as the distribution of model classes. Linear and tree-based models are the most common models. **D) Sample size of analyzed cohorts**. While some papers featured large patient cohorts ranging from 2,000 to 20,000 patients, the majority of papers examined smaller patient populations. Abbreviations: AIS; ASIA Impairment Scale, CSF; Cerebrospinal fluid, FIM; Functional Independence Measure, GRASSP; Graded Redefined Assessment of Strength, Sensation, and Prehension, ISNCSCI; International Standards for Neurological Classification of SCI, MP; motor point, MRC; Medical Research Council manual muscle testing, MRI; magnetic resonance imaging, NLI; neurological level of injury, SCIM III; Spinal Cord Independence Measure III, UEMS/LEMS/TMS; upper extremity motor score/lower extremity motor score/total motor score.

In line with the focus on computational architectures comprising limited parameterization, the majority (74.2%) of studies were conducted in small patient cohorts of less than 500 subjects. Despite the large heterogeneity expected in SCI patients, few studies [15, 33, 42, 62] used larger sample sizes beyond 2000 subjects. It is worth noting the study by Kapoor D., et al. [33], comprising 20790 individuals. It bench-marked different machine learning models (ridge classifier, support vector machine, elastic net, logistic regression, ensemble model, convolutional neural network, random forest, and naive Bayes) on data from the National Spinal Cord Injury Statistical Center (NSCISC) database to predict AIS scores at hospital discharge.

### Qualitative assessment of studies

We summarize the qualitative results obtained from scoring different aspects relevant to a data-driven analysis in Figure 3; individual scores are provided in Table S4.

**Fig. 3.**
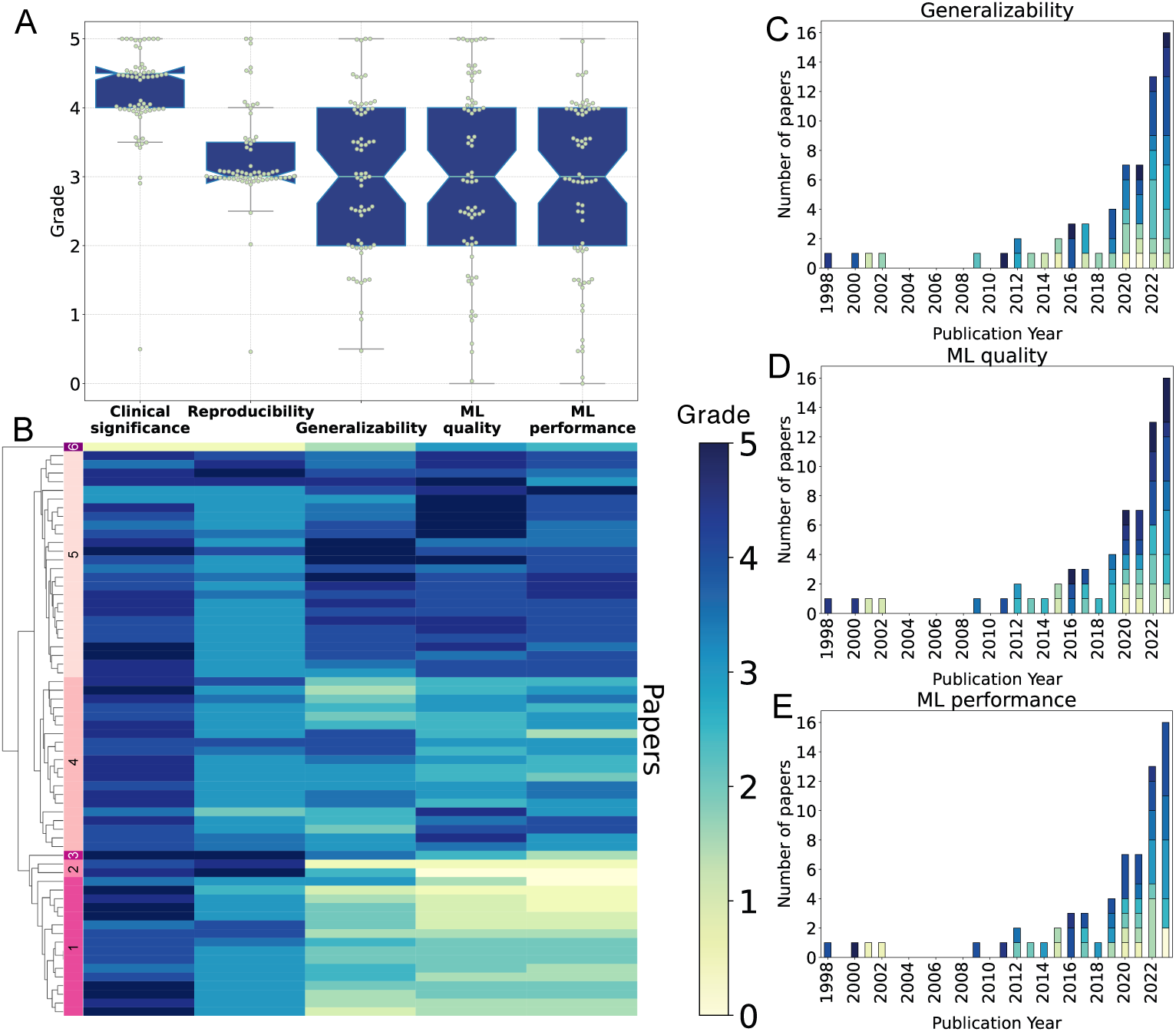
Overview of the grading of clinical relevance and machine learning categories of the included articles. **A) Scoring performance overview** stratified by the five assessment categories. Scores between 0 to 5 with intervals of 0.5 are possible. The boxplot illustrates the median (light blue line) and quartile ranges. The whiskers extend to the farthest data point within 1.5x of the relevant interquartile range. Individual scores are shown with jitter. **B) Heatmap overview of the scores** achieved by each study (row) subject to hierarchical clustering as visualized by the dendrogram. The hierarchy is flattened into 6 clusters indicated by color. **C-E) Visualization of scores as a function of time** for the generalizability (C), machine learning quality (D), and machine learning performance (E)

We observe that the majority of papers scored consistently high in clinical significance (median 4.5, minimum 3.5 (excluding three outliers), Figure 3A). The outliers, scoring 0.5, 3 and 3 [19, 37, 40], were computationally focused but lacked a clear definition of the clinical problem they were addressing.

A total of 83% of the papers achieved a *Reproducibility* score of at most 3.5 (Figure 3A). Higher scores (*>* 3.5) were predominantly related to making the data available (on request) (n=11) and/or the code base available (n=4). Three percent of the studies did not provide access to code and had unclear definitions of the model inputs and outcomes, resulting in a reproducibility score below 2.5.

For the other evaluation criteria (*Generalizability*, *ML Quality*, and *Performance*), we observed varied results across the included articles, with a median score of 3 in all categories, reflecting a broad range of scores (Figure 3A). Many studies (n = 34) addressed the problem of generalization by either external validation or performing a clean train-test data split.

Figure 3B groups all 66 studies using hierarchical clustering. Except for one study [40] that failed to meet both *Clinical Significance* and model *Reproducibility* (cluster 6) and one study that had great *Clinical Significance* and *Reproducubility* but lower grades in the rest of the categories (cluster 3), our clustering revealed four sets of comparable study designs: i) those achieving adequate (score above 3) ratings across all criteria (n=26, cluster 5), ii) those yielding intermediate (*≈* 3) model *Generalizability*, *ML Quality* and *ML Performance* (n = 20, cluster 4), iii) those failing to address the latter three criteria (n = 16, *Generalizability*, *ML Quality* and *ML Performance* below 1, cluster 1), and iv) studies of poor generalizability and ML quality but having good *Reproducibility* (cluster 2).

Regarding the longitudinal development across publication years (Figure 3C-E), we did not observe a clear trend of quality improvement despite the noticeable increase in the number of publications focusing on applying data-driven approaches to SCI-related prediction tasks (Figure 3D).

## Discussion

Our literature review identified 66 studies addressing the topic of data-driven prediction of recovery after traumatic SCI. These studies employed a range of computational models including linear regression, tree-based models, and neural networks. The choice of model architectures appeared largely influenced by data constraints and the need for reproducible and interpretable implementations inherent in standard machine learning models. Still, a recent trend toward more complex approaches is observed. Data availability also strongly influenced the choice of input features and outcome variables. Predominantly, easily accessible data such as patient demographics and neurological assessments were favored in prediction models. Studies incorporating imaging, functional (e.g. walking ability), molecular, or hematological data as input features were less common. The most commonly used outcomes were AIS grades, walking ability, or SCIM III. Despite the acknowledged importance of sexual, bowel, and bladder functions for individuals living with SCI [86], these endpoints are understudied, likely due to the lack of available data on these specific aspects. Whereas data on sexual function is particularly limited, bladder and bowel functions are part of the SCIM III assessment. The SCIM III is frequently assessed as part of SCI clinical trials [87–90] and observational cohorts [5–7, 81].

A striking observation was the notable variability and frequently inadequate precision in defining and reporting time points for input features (up to approximately 56 days) and outcome variables (typically around 1 year). The choice of outcome time-point (*≈* 1 year) is clinically reasonable, considering the limited remaining recovery potential beyond this timeframe [91].

However, our evaluation also highlights the necessity for improved reporting practices to enhance reproducibility and applicability. In recent decades, the field has established several large-scale observational data registries (e.g. EMSCI, RHSCIR) that address these gaps.

The qualitative assessment revealed consistently high *Clinical significance* scores across all articles, indicating that SCI recovery prediction is driven by clinical application, facilitated by close collaboration between clinical data providers and computational researchers. In contrast, the *Reproducibility* was observed to be relatively low, leaving ample room for improvement. Specifically, while input, output, and chosen models were reported, the provision of code — which is essential for study reproducibility — was rare, yet indispensable upon the publication of data-driven models [92]. We also observed substantial variability in the scoring of *Generalizability*. In general, the importance of *Generalizability* is acknowledged in the field, however, its implementation is often suboptimal. While employing an external validation set would be ideal, adopting a rigorous train-test split can serve as an initial step toward more genuinely externally validated approaches.

In summary, there is a noticeable surge in interest in data-driven SCI recovery predictions, which has inspired a range of promising predictive studies of high clinical relevance. As this field evolves, it is crucial to effectively utilize and report datadriven prediction models to maximize their potential for clinical deployment. Based on our quality assessment, there is room for improvement in terms of reporting and defining outcome variables, as well as ensuring reproducibility and generalizability of the employed models. In the following, we outline possible future directions and key challenges, mention parallels to other clinical applications regarding data types, and outline aspects of promising model architectures and training regimes.

### Future directions and key challenges of data-driven recovery prediction

Data availability and diversity remain a limitation for data-driven predictions of SCI recovery. Data-efficient training strategies and optimal use of the available data address this challenge. Missing and incomplete data, particularly in a longitudinal context, is common in biomedical data science. In the SCI field, missing data predominantly arise due to patient transfers between treatment and rehabilitation facilities, comorbidities, and patient compliance with elaborate assessment protocols such as SCIM and ISNCSCI [93]. It is widely accepted to rely on complete-case analysis rather than imputed data [93–95]. Carefully applied imputation could, however, increase the pool of training samples and lead to more robust predictions. Bourguignon et al. provide a comprehensive evaluation of diverse imputation strategies in the SCI domain to achieve this [93]. Importantly, depending on the target data type and clinical context, different imputation strategies should be applied (e.g. ordinal imputation [96], last observation carried forward [97], or multiple imputation [98]). In addition to maximizing data usage, strategies from limited data learning, such as auxiliary task learning (training for a subtask that is indirectly linked to the objective of interest) [99], self-supervision (contrasting sample subgroups) [100] and data augmentation can be harnessed.

Traditional data augmentation aims to enhance the performance through various transformations, such as geometric transformations and color space augmentations in imaging [101], aiming to increase the diversity and quantity of the data. Augmenting datasets with limited labels using semi-supervised [102] and active learning [103] could further improve model performance through strategic data labeling and the incorporation of unlabeled datasets [104, 105]. Moreover, data from an external domain might prove useful in settings with limited data. Transfer learning is a compelling strategy that leverages a large dataset from a source domain to (pre-)fit model weights, leading to improved performance upon the target domain upon model refinement. This approach has previously proven successful in related applications such as neurodegenerative disorders [106]. While general data sets are useful, data that are related to the target task (e.g. same data type, same anatomical context) would be preferable [107]. While ISNCSCI data may be unique, parallels for spinal imaging and (multi-) omics data in SCI-related tasks may be readily accessible.

A common limitation to increasing sample size may be data privacy concerns restricting large-scale data centralization. Federated learning represents an alternative way of collaborative and privacy-preserving multisource data digestion [108]. This is especially relevant in scenarios where direct data sharing among institutions is impractical [109]. In the SCI recovery prediction field, techniques for limited data learning combined with a federated learning framework hold the potential to maximize the usage of any data available. This would allow exploration of advanced architectures, beyond the current standard linear and tree-based approaches.

Currently, prediction models are predominantly based on readily available data types (ISNCSCI assessments, image features, demographics). Using underexploited data types like omics and images could improve model predictions. Although the collection and curation of these data types may be challenging, they hold the potential to uncover a deeper biomedical understanding of the physiological processes underpinning SCI recovery in addition to identifying predictive biomarkers. Similarly, combinations of more established data types with imaging and molecular characterization of CSF or blood could be considered. Multi-modal learning that harnesses diverse molecular data alongside clinical information could offer nuanced insight into patient recovery trajectories [110]. Independent of the specific data types, innovative architectures like graph neural networks (GNNs) [111], attention mechanisms [112], and transformers [113] could pave the way for next-generation modeling of spinal structures and patient recovery patterns. These architectures have previously been shown to effectively interpret complex data structures and longitudinal settings [114–116].

As models become more intricate, interpretability and explainability become more critical. Model transparency is essential for their acceptance as clinical decision support [12, 117, 118]. This is addressed in the topic of causal inference which enables tracing the prediction formulation [119]. The development of a data-driven prediction pipeline involves several critical steps including data collection, preprocessing, hyperparameter tuning, and model evaluation, ideally in collaboration with clinical experts. Automating (part of) this pipeline could accelerate the development and validation of solutions. AutoML (Automated Machine Learning) [120] is one example of such a streamlined pipeline, simplifying model deployment by automating tasks such as model selection and tuning, making it easier and faster to integrate with clinical applications.

In our current review, none of the included studies effectively accounted for the time dependence of SCI recovery beyond the input data and output label inclusion brackets. SCI recovery is a process. The longitudinal character of available SCI data may hold information regarding the specific recovery trajectory, implying that data from repeated assessments should be considered [121]. Going even further in temporal data resolution, sensor data from technologies such as wearables [122] and exoskeletons [123] are available for real-time monitoring of SCI patients. As data-driven models have previously been used to recognize physical activity in patients with SCI [124, 125], these could further enhance recovery prediction.

Finally, to ensure optimal training and objective performance evaluation, the SCI community needs to agree on standards regarding recovery prediction performance evaluation and endpoints of interest. In this review, we saw a variety of tasks associated with SCI recovery. While ISNCSCI composite scores, AIS grade conversion, and SCIM items are widely applied, the clinical relevance, objectivity, and prediction suitability need to be assessed. Ceiling and flooring effects [31], as well as co-linearity of contributing sub-scores, are common limitations in this context [126]. To enhance the understanding and accuracy of SCI recovery evaluations, it is crucial to account for the predominantly non-decreasing nature of patient recovery trajectories (i.e. patients generally exhibit improvement over time). Furthermore, when assessing ordinal scale labels like SCIM or AIS grades, metrics and loss functions suitable for these data types (i.e. ordinal) should be the standard [127]. Stringent data splits are equally important. To demonstrate model generalizability and clinical relevance, even given small sample sizes, (leave-one-out) cross-validation [128] and external test sets are key. In summary, we propose that by embracing state-of-the-art developments in machine learning, beyond standard architectures and training regimes, new insights into spinal cord injury can be discovered. This will pave the way towards robust and clinically relevant predictions.

## Conclusion

Data-driven approaches from the realm of machine and deep learning hold promise to improve the prediction of recovery outcomes in SCI. The use of a variety of data types and implementation architectures reflects the growing trend in the SCI research community to harness these approaches in a clinical context. However, accomplishing clinical translation requires careful consideration of model validity, causality, and practical deployability, in addition to growing and maintaining increasingly detailed databases. Currently, data-driven SCI recovery prediction models often rely on standard architectures applied to comparably small sample sizes. This motivates further investigation of modern training paradigms and architectures that embrace the challenges in SCI recovery, such as limited data and model robustness. Data-driven predictions have the potential to inform rehabilitation strategies, enhance the design of clinical trials, and consequently, enhance the overall quality of life for individuals affected by SCI.

## Data Availability

All data produced in the present study are available upon reasonable request to the authors

## Acknowledgements

This study was supported by the Swiss National Science Foundation (Ambizione Grant, #PZ00P3 186101), Wings for Life Research Foundation (#301), Marie Sk-lodowska-Curie Actions (ReWIRE, #101073374), and the International Foundation for Research in Paraplegia (IRP, P192). SB was supported by the Botnar Research Centre for Child Health Postdoctoral Excellence Programme (#PEP-2021-1008). The funders did not specify the study design, data collection, analysis, or the decision to publish and preparation of the manuscript.

During the preparation of this work, the author(s) used ChatGPT-4 in order to extract information from papers. After using this tool/service, the author(s) reviewed and edited the content as needed and take(s) full responsibility for the content of the publication.

## Supplementary

Order of labeling for study exclusion:

**Table S1.** Description of hierarchical exclusion criteria.

| Item | Description |
| --- | --- |
| 1 | Review, collection or commentary |
| 2 | Non-human |
| 3 | Not traumatic (or ischemic) spinal cord injury, such as stroke, degenerative (e.g. myelopathy), pediatric |
| 4 | No prediction model or validation study of a previously described model (including linear regression models fitted to new data but excluding linear regression models with parameters from a previous paper.) |
| 5 | No recovery prediction |
| 6 | Different outcome measure |

**Table S2:**
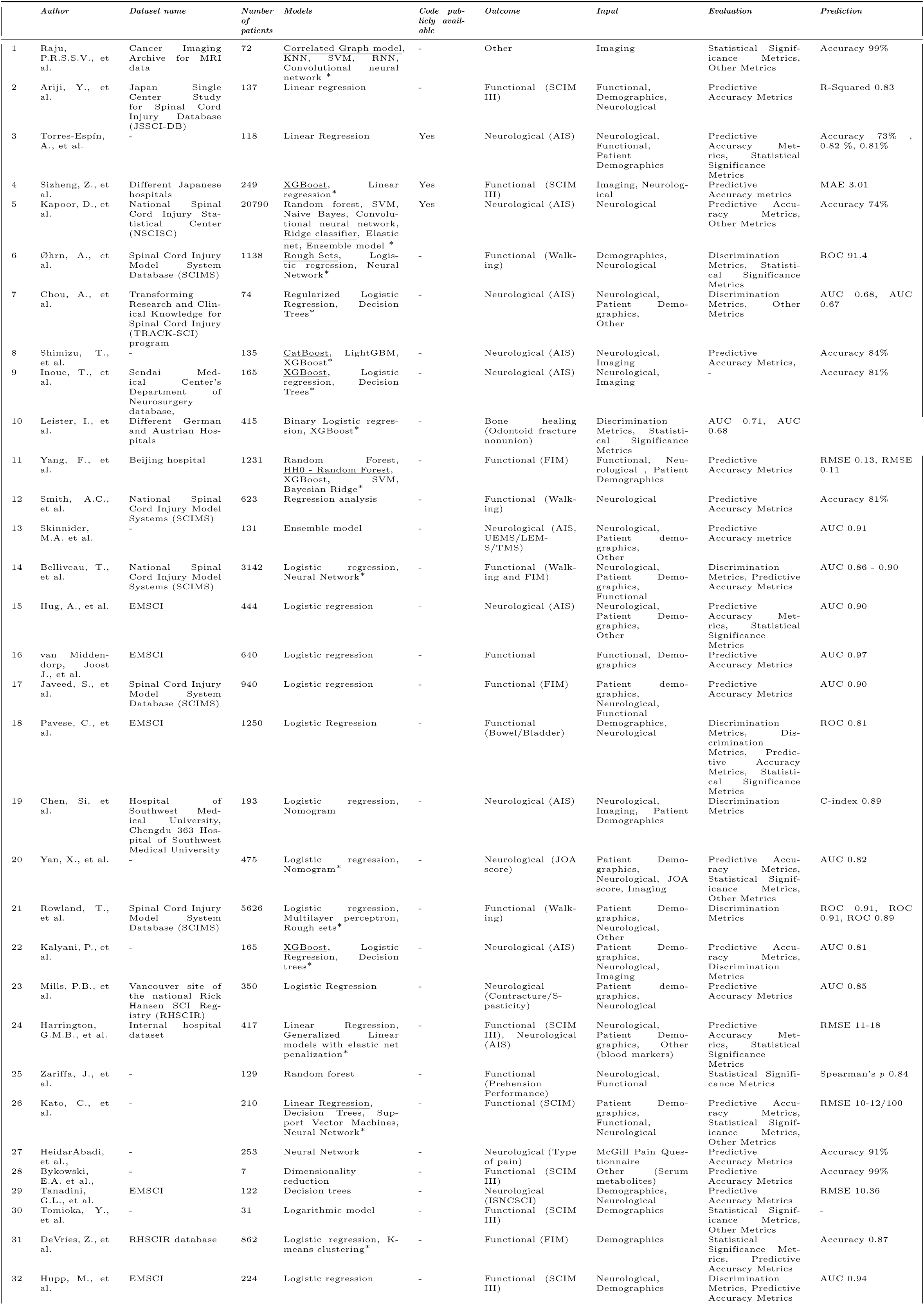
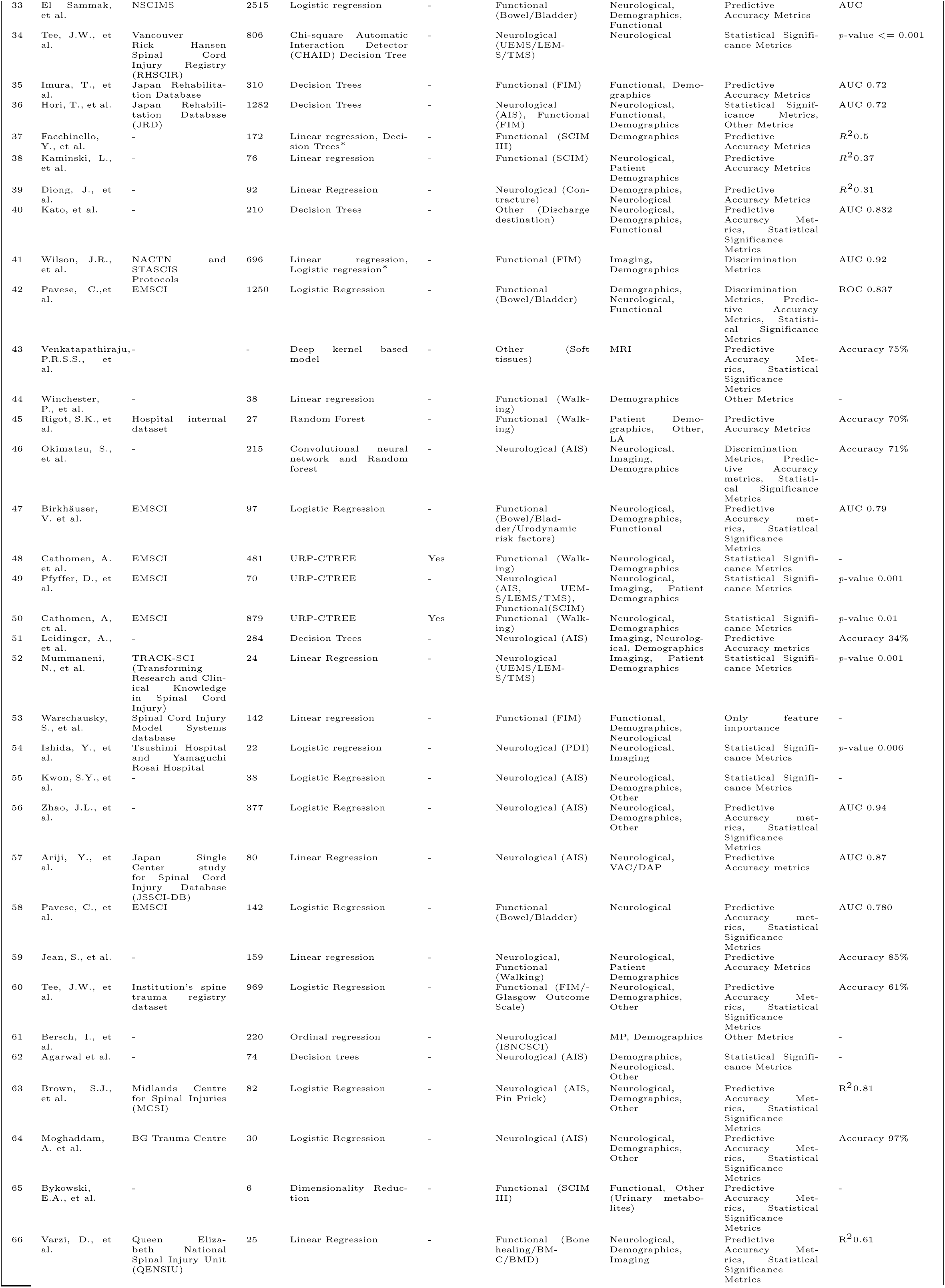
Overview of the information retrieval from the systematic review. The order of articles is the same as in the heatmap in Figure 3B.

**Table S3.**
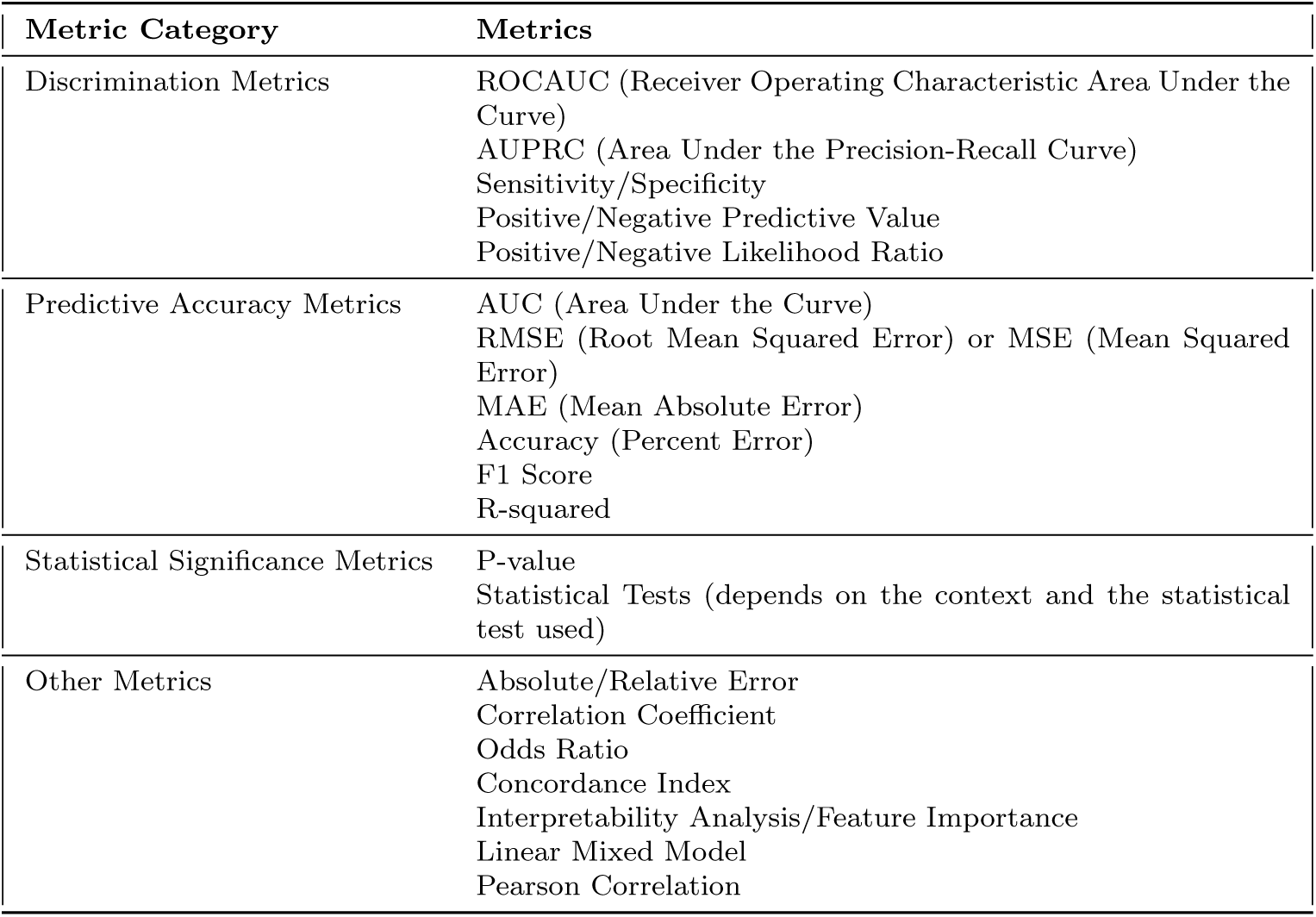
Evaluation metrics by category.

**Table S4:** Grades of the studies in the systematic search on the scale 0-5 using the mean of two scores (two raters). The order of articles is the same as in the heatmap in Figure 3B.

|  |  | *Clinical*<br>*significance* | *Reproducibility* | *Generalizability* | *ML quality* | *ML performance* |
| --- | --- | --- | --- | --- | --- | --- |
| 1 | Raju, P.R.S.S.V., et al. | 0.5 | 0.5 | 1.5 | 3 | 2.5 |
| 2 | Ariji, Y., et al. | 4.5 | 4 | 3.5 | 4.5 | 4 |
| 3 | Torres-Espín, A., et al. | 3.5 | 4.5 | 3.5 | 4.5 | 4 |
| 4 | Sizheng, Z., et al. | 4.5 | 5 | 4 | 4 | 3.5 |
| 5 | Kapoor, D., et al. | 4.5 | 4.5 | 4.5 | 5 | 3 |
| 6 | Øhrn, A., et al. | 3 | 3 | 4 | 4.5 | 5 |
| 7 | Chou, A., et al. | 3 | 3 | 3 | 5 | 4 |
| 8 | Shimizu, T., et al. | 4.5 | 3 | 3.5 | 5 | 4 |
| 9 | Inoue, T., et al. | 4 | 3 | 3.5 | 5 | 4 |
| 10 | Leister, I., et al. | 3.5 | 3 | 4 | 5 | 3.5 |
| 11 | Yang, F., et al. | 4 | 3.5 | 4 | 5 | 3.5 |
| 12 | Smith, A.C., et al. | 4.5 | 3 | 5 | 3.5 | 3.5 |
| 13 | Skinnider, M.A. et al. | 5 | 4 | 5 | 4 | 4 |
| 14 | Belliveau, T., et al. | 4 | 3 | 5 | 5 | 4 |
| 15 | Hug, A., et al. | 3.5 | 3 | 4 | 3.5 | 4 |
| 16 | van Middendorp, Joost J., et al. | 4 | 3.5 | 5 | 4 | 4.5 |
| 17 | Javeed, S., et al. | 4 | 3 | 4.5 | 4 | 4.5 |
| 18 | Pavese, C., et al. | 4.5 | 3.5 | 4 | 4 | 4.5 |
| 19 | Chen, Si, et al. | 4.5 | 3 | 4.5 | 4 | 4 |
| 20 | Yan, X., et al. | 4.5 | 3 | 4 | 4 | 4 |
| 21 | Rowland, T., et al. | 4 | 3 | 4.5 | 4.5 | 4 |
| 22 | Kalyani, P., et al. | 4 | 3 | 4 | 4.5 | 4 |
| 23 | Mills, P.B., et al. | 4 | 3 | 4 | 4 | 4 |
| 24 | Harrington, G.M.B., et al. | 5 | 3 | 4 | 4.5 | 3.5 |
| 25 | Zariffa, J., et al. | 5 | 3 | 4 | 3.5 | 4 |
| 26 | Kato, C., et al. | 4.5 | 3 | 3.5 | 4 | 4 |
| 27 | HeidarAbadi, et al., | 4.5 | 3 | 3 | 4 | 4 |
| 28 | Bykowski, E.A. et al. | 4.5 | 4 | 2 | 2.5 | 2.5 |
| 29 | Tanadini, G.L., et al. | 5 | 3 | 1.5 | 2.5 | 3 |
| 30 | Tomioka, Y., et al. | 4.5 | 3.5 | 2 | 3 | 3.5 |
| 31 | DeVries, Z., et al. | 4 | 3 | 2.5 | 3 | 2.5 |
| 32 | Hupp, M., et al. | 4 | 3 | 2 | 2.5 | 3 |
| 33 | El Sammak, et al. | 4.5 | 3 | 2.5 | 2.5 | 3 |
| 34 | Tee, J.W., et al. | 4.5 | 3 | 4 | 2.5 | 1.5 |
| 35 | Imura, T., et al. | 4 | 4 | 4 | 3 | 3 |
| 36 | Hori, T., et al. | 4 | 3.5 | 4 | 2.5 | 3 |
| 37 | Facchinello, Y., et al. | 4.5 | 3 | 3.5 | 3 | 2.5 |
| 38 | Kaminski, L., et al | 4.5 | 3 | 3 | 2.5 | 2.5 |
| 39 | Diong, J., et al. | 4.5 | 3 | 3 | 2.5 | 2 |
| 40 | Kato, et al. | 4 | 3 | 3 | 3 | 3.5 |
| 41 | Wilson, J.R., et al. | 4.5 | 3 | 3.5 | 3 | 3.5 |
| 42 | Pavese, C.,et al. | 4.5 | 3 | 3.5 | 2.5 | 3 |
| 43 | Venkatapathiraju, P.R.S.S., et al. | 3.5 | 2 | 2.5 | 4.5 | 3 |
| 44 | Winchester, P., et al. | 4.5 | 3 | 2.5 | 3.5 | 4 |
| 45 | Rigot, S.K., et al. | 4 | 3 | 2 | 4 | 4 |
| 46 | Okimatsu, S., et al. | 4 | 3.5 | 3 | 4.5 | 3 |
| 47 | Birkhäuser, V. et al. | 4 | 3.5 | 3 | 3.5 | 3 |
| 48 | Cathomen, A. et al. | 5 | 5 | 3.5 | 2.5 | 1.5 |
| 49 | Pfyffer, D., et al. | 4 | 4.5 | 0.5 | 0.5 | 0.5 |
| 50 | Cathomen, A, et al. | 4.5 | 5 | 2.5 | 0 | 0 |
| 51 | Leidinger, A., et al. | 3.5 | 3 | 3 | 1.5 | 0 |
| 52 | Mummaneni, N., et al. | 5 | 2.5 | 1 | 0.5 | 0.5 |
| 53 | Warschausky, S., et al. | 4.5 | 3 | 1.5 | 1 | 0.5 |
| 54 | Ishida, Y., et al. | 5 | 3 | 2 | 1 | 0.5 |
| 55 | Kwon, S.Y., et al. | 5 | 3 | 2 | 1 | 1 |
| 56 | Zhao, J.L., et al. | 3.5 | 4 | 2 | 1 | 1 |
| 57 | Ariji, Y., et al. | 4 | 4 | 1.5 | 1.5 | 1.5 |
| 58 | Pavese, C., et al. | 4 | 3.5 | 2.5 | 2 | 2 |
| 59 | Jean, S., et al. | 4 | 3 | 2 | 2 | 2 |
| 60 | Tee, J.W., et al. | 4 | 3 | 2 | 2 | 2 |
| 61 | Bersch, I., et al. | 3.5 | 3 | 2.5 | 2 | 1.5 |
| 62 | Agarwal et al. | 4 | 3 | 2.5 | 1.5 | 1.5 |
| 63 | Brown, S.J., et al. | 5 | 3 | 2 | 2 | 2 |
| 64 | Moghaddam, A. et al. | 5 | 3 | 1.5 | 2 | 2 |
| 65 | Bykowski, E.A., et al. | 4.5 | 3 | 1.5 | 1.5 | 1.5 |
| 66 | Varzi, D., et al. | 5 | 3 | 1 | 1.5 | 1.5 |

